# Community-based emergency transport system in Shinyanga, Tanzania: a local innovation to avert maternal and newborns deaths for a low-resource setting

**DOI:** 10.1101/2022.12.20.22283749

**Authors:** Castory Munishi, Gilbert Mateshi, Linda B. Mlunde, Belinda J Njiro, Jackline E Ngowi, James T. Kengia, Ntuli A. Kapologwe, Linda Deng, Alice Timbrell, Wilson Kitinya, Andrea B. Pembe, Bruno F. Sunguya

**Affiliations:** Muhimbili University of Health and Allied Sciences, Dar es Salaam, Tanzania; President’s Office Regional Administration and Local Government, Dodoma, Tanzania; Touch Foundation, Mwanza, Tanzania

**Keywords:** Maternal mortality, emergency referral, perinatal mortality, cost effectiveness

## Abstract

**Introduction:** In achieving sustainable development goal 3.1, Tanzania needs substantial investment to address the persistent burden of maternal mortality. Efforts are needed to curtail the three delays which underlie most of the deaths. The government of Tanzania implemented a community-based emergency transport intervention to address the second delay through m-mama program to address these delays. This examined the cost-effectiveness of this intervention as compared to the use of a standard ambulance system alone.

**Materials and Methods:** Program data provided by the program implementers was used in this secondary data analysis. The data included costs of referral services using the Emergency Transportation System (EmTS) and comparing with the standard ambulance system. Analysis was conducted using Microsoft Excel to generate data that was fed in a TreeAge Pro Healthcare 2022 model. The cost and effectiveness data were discounted at 5% to make fair comparisons of the two systems.

**Results:** A total of 989 referrals were made where, 30.1% were completed using the standard referral system using ambulance, and the majority (69.9%) were completed using the EmTS. The Emergency transport system costed USD 170.4 per completed referral compared to USD 472 per referral using ambulance system alone in the six councils of Shinyanga region where the intervention was conducted.

**Conclusion:** The introduction of m-mama emergency transportation system has proved to be cost effective compared to the use of standard ambulance system alone in Shinyanga region. Given the high burden of maternal mortality in Tanzania, this community-based program can save implementation costs and may be more sustainable should be scaled up to other regions in Tanzania. Using implementation science methods while scaling up, lessons learned in various regions may further improve the effectiveness of the EmTS.

## Introduction

The first target of Sustainable Development Goal number three aims to reduce global maternal mortality ratio (MMR) to less than 70 per 100,000 live births by 2030 [1]. With a few years remaining towards 2030, countries in sub-Sahara need innovative approaches in addressing maternal mortality. Like other countries in the region, Tanzania has one of the highest MMR in the world [2,3]. Most of the maternal deaths are preventable, it is therefore possible to cut down MMR using local innovations and evidence generated within the same context [4].

Causes of maternal and perinatal deaths in Tanzania fit in a three-delay model [5]. The first, is the delay in making the decision to seek care when experiencing an obstetric emergency; the second, is the delay in reaching an appropriate obstetric facility once the decision has been made to go; and third, is the delay in receiving adequate and appropriate care once the facility has been reached. These delays do not operate independently [6]. For example, expectations of transport delays or of low-quality care at the nearest facility or the distance to reach the facility with no appropriate means to reach there, influence the initial decision to seek care.

Community level interventions have an important role to play in addressing the burden of maternal mortality [7]. Such interventions include training for community health workers to recognize danger signs in pregnancy, provision of nutritional supplements to pregnant women, increasing community awareness of danger signs of pregnancy, and need to seek early care [8,9]. Community based interventions have also proven useful in addressing the three delays [10].

The m-Mama program was designed and implemented by the government of Tanzania in collaboration with Touch Foundation, Pathfinder International, and Vodafone foundation in Shinyanga region, Tanzania. In this program, an innovative Emergency Transportation System (EmTS) was designed to utilize a digital technology application that could remotely triage patients and dispatch an ambulance or community-based drivers to transfer the patient to a designated health facility. While directly addressing transportation, the program also included a focus on health system strengthening and community education initiatives through Community Care Groups to encourage women to seek healthcare and ensure women receive high-quality care upon reaching the appropriate health facility. Program evaluation demonstrated a decline in maternal mortality following the introduction of this program. However, cost-effectiveness of this intervention was not known. This study therefore aimed to determine the cost-effectiveness of this intervention compared with the standard emergency referral system.

## Methods

### Study design and settings

This cost-effectiveness analysis was part of the endline m-mama program evaluation and used secondary data from the program documents. The analysis aimed to compare the effectiveness of the emergency transport system (EMT) for pregnant women in the Shinyanga region. The emergency transport system is comprised of normal ambulance services and a community taxi when an ambulance isn’t available. For this analysis, the EMT is compared against the standard model of referral which is the normal ambulance transport system with no community tax backup. The EmTS was used in the Shinyanga region to provide an improved means of transport for pregnant women and their newborns during times of emergencies in six councils.

The m-mama program introduced dispatchers to the council level. A toll-free number in case of emergency to the dispatch center was also introduced to offer a 24/7 access. Upon receiving an emergency call, a trained dispatcher would remotely triage the patient after an initial assessment through a series of pre-defined questions. Supported by a decision-making application running on a tablet, the dispatcher would enter the information received, and the application trigger a set of sequential logical questions. The application would then indicate whether the patient need to be transferred to a designated health facility based on her condition. This process would take an average of two minutes.

The dispatch application is integrated with a mobile payment system ensuring a fast and responsive payment for a community driver who completed the referral transport action. To ensure sustainability the program followed a progressive transition on its funding approach, at the start (first stage January to December 2019) the program financed 80% of the transportation costs, on the second stage (January to December 2020) the program financed 50% and the government 50% and on third stage from January 2021 the government financed 100% of the EmTS.

### Target Population

The emergency transportation system was built to facilitate transport of women during emergencies from the community to facilities, or from one facility to another in Shinyanga Region. The drivers who participated in the program were community members who owned vehicles and were willing to work in EmTS. They were trained and enrolled and rendered ready to handle the emergencies. The drivers in the standard ambulance system were professional drivers employed and deployed to government facilities in the region.

### Source of data

Costs data for the emergency transportation system and the use of ambulances were provided by the m-Mama program. The data covered the costs incurred in the final year spanning July 1^st^, 2020, to June 30^th^, 2021.

### Variables and Analysis Context

The key variables and parameters in the cost effectiveness analysis were perspective in which a provider’s perspective was used whereby costs incurred by the providers during the provision of EmTS were compared. During the analysis, cost of emergency transportation system using community taxis was compared against the standard ambulance system. The evaluated costs and outcomes covered July 1^st^, 2020, to June 30^th^ 2020. A 5% discount rate used was provided by the Bank of Tanzania. Outcome measure for effectiveness was the number of completed referrals. The costs in the analysis were reported in Tanzanian shillings and they were reported for July 2020/June 2021 a twelve monthes period and these were further converted to USD using the BoT exchange rates at the time of implementation. The model used for analysis was a simple decision tree to determine the effectiveness of the two interventions.

The following assumptions were considered in the analysis that the EMT complements the standard of care (Ambulance only) therefore it was assumed to be the standard of care plus the community taxi services. Second, the cost of service included all the costs incurred by the provider/ region when providing referral services and it excludes the diesel cost. The final cost was then the services cost plus the diesel cost. Third, this analysis assumed that all the referrals were completed during the same time the costs were incurred (July 2020/June 2021). Lastly, all completed referrals were effective referrals and the decision tree for both strategies wasn’t branched into effective and ineffective referrals.

Data was cleaned using Microsoft excel 2016 version, the total costs were then calculated and summarized in tabular format and then inputted into a TreeAge Pro Healthcare 2022 software using a decision tree model for analysis of costs and effectiveness. The decision tree model generated a cost-effectiveness ranking report. Finally, the costs of both ambulances and the new emergency transportation were discounted at 5% as to make fair comparisons as costs and outcomes did not occur at the same time [11].

The following calculations were made during the analysis: the Incremental cost (Incr Cost) which was calculated as the difference in cost between the EMT and the standard referral system. Incremental Effectiveness (Incr Eff) was calculated as the difference between the effectiveness (Outcome) of the EMT and the standard referral system. Cost-effectiveness (CE) was calculated as the ratio of cost and effectiveness (Outcomes) for each intervention and Incremental cost-effectiveness ratio (ICER) was calculated as the ratio of incremental cost to incremental effectiveness. This represents the additional cost per unit of effectiveness to switch to a more effective strategy

### Ethical consideration

Ethical approval for this evaluation was granted by the Muhimbili University of Health and Allied Sciences Institutional Review Board (MUHAS-REC-11-2021-885). Permission to collect data from Shinyanga region was granted by the President’s Office Regional and Local Government Authority and the office of the Regional Medical Officer in Shinyanga. Written informed consent was obtained from participants before data collection. Privacy and confidentiality were maintained. The data obtained from this program evaluation were kept as strictly confidential, were accessible to only the named investigators and has been stored on password-protected computers.

## Results

### General characteristics of emergency referrals

Data on costs and completed referrals for the Shinyanga region, from July 2020 to June 2021 (FY 2020/21) were used for this cost-effectiveness analysis. During this period, the six councils completed a total of 989 referrals. These were completed using both the standard ambulance referral services and the community taxis. Out of all referrals, 30.1% were completed using the standard referral system using ambulance services while the majority 69.9% were completed by the use community taxis; an intervention that was introduced along the standard referral system.

In Shinyanga municipal council majority (90%) of the referrals were carried out using ambulances. Msalala, Kishapu, and Shinyanga district council which are more rural areas had a higher proportion of trips completed through community taxis (**Table 1)**.

**Table 1.** Total number of referrals completed.

| Council | Ambulance referrals, n (%) | Community taxi referrals, n (%) |
| --- | --- | --- |
| Kishapu | 51 (17.6) | 239 (82.4) |
| Kahama | 23 (43.4) | 30 (56.6) |
| Msalala | 15 (13.3) | 98 (86.7) |
| Shinyanga DC | 64 (19.3) | 267 (80.7) |
| Shinyanga MC | 117 (90.0) | 13 (10.0) |
| Ushetu | 28 (38.9) | 44 (61.1) |
| **Total** | **298 (30.1)** | **691 (69.9)** |

### Total Cost of emergency referrals

A total of ZS 389.4 million was spent during the FY2020/21 among six councils of Shinyanga region while completing all the referrals. Most of the cost was incurred on ambulance trips even though there were just a few as seen in **Table 2**. The standard ambulance referrals represented 83% of the total cost used, with Kishapu covering most of the expenditures at 29% and Shinyanga MC being the least at 7%. For community taxi referrals, the highest cost came from Kishapu (40%), and the least was from Shinyanga MC (1%). Table 2. Shows the cost of referrals between councils for both ambulance referrals and community taxis.

**Table 2.** Cost of referrals completed for FY2020/21.

| **Council** | **Cost of Ambulance referrals (TZS)** | **Cost of Community taxi referrals (TZS)** |
| --- | --- | --- |
| Kishapu | 94,000,000 | 26,000,000 |
| Kahama | 73,000,000 | 2,440,000 |
| Msalala | 40,000,000 | 8,186,000 |
| Shinyanga DC | 36,000,000 | 23,758,000 |
| Shinyanga MC | 23,000,000 | 374,000 |
| Ushetu | 59,000,000 | 3,594,000 |
| **Total** | **325,000,000** | **64,352,000** |

### Cost-effectiveness

When calculating the cost-effectiveness, all referrals were considered completed referrals. Table 3 shows the undiscounted cost and effectiveness between the standard intervention (ambulance only) and the new intervention (ambulance plus community taxis for FY2020/21.

**Table 3.** Summary of Undiscounted cost and effectiveness for Shinyanga region.

| **Interventions/Strategy** | **Costs in TZS** | **Cost in USD\*\*** | **Number of referrals** |
| --- | --- | --- | --- |
| **Standard (Ambulance only)** | 325,000,000 | 140,715.18 | 298 |
| **EMT (Ambulance+ Community Tax)** | 389,352,000 | 168,577.65 | 989 |

*\*Note: the exchange rate used =2,309.63 (BoT, 20 Feb 2022), \*\*USD = United States Dollar*

Using the TreeAge Pro Healthcare 2022, the incremental cost and effectiveness between the standard and the new intervention were $27,862 and $ 657 respectively and an incremental cost-effectiveness ratio (ICER) of $40.3 as seen in Table 4. The cost-effectiveness ratio for the standard and the new intervention was $472.2 and $170.4 respectively per referral completed. This shows that the EmTS is more cost-effective when compared to the standard as it gives a lower cost-effectiveness ratio. Finally, both the costs and effectiveness were discounted at 5%. The total discounted costs for standard and new interventions were $133,679.4 and $160,148.8 respectively. The discounted effectiveness was $283 and $940 for standard and new interventions respectively.

**Table 4:** Cost-effectiveness ranking.

| **Cost-Effectiveness Rankings Report** |  |  |  |  |  |  |  |
| --- | --- | --- | --- | --- | --- | --- | --- |
| **Strategy** | **Cost (\$)** | **Incr Cost\*** | **Eff\*\*** | **Incr Eff** | **C/E** | **ICER** | |
| Standard referral system (Ambulance only) | 140,715 | 27,862 | 298 |  | 472.2 |  |  |
| Emergency Transport System (Ambulance plus community Taxi) | 168,578 | 657 | 989 | 691 | 170.4 | 40.3 |  |

*Incr Cost: Incremental Cost; Eff: number of completed referrals; Incr Eff: Incremental cost-effectiveness; C/E: Cost-effectiveness ratio; ICER: Incremental cost-effectiveness ratio*

## Discussion

The Emergency Transport System of the m-mama program was a more cost-effective intervention in addressing emergency referrals compared to the standard emergency referral system in Shinyanga region. This innovative system costs $170.4 per completed referral as compared to $472 per referral by using ambulances alone. The program data revealed that, the emergency referral system was used more in the rural areas where ambulances have limited functionality given the remoteness of the areas.

Comparing the findings of this evaluation to an emergency referral system using ambulances that were implemented in Burundi by Médecins sans Frontières (MSF) reported the cost per referral to be 61£ [12] equivalent to $121 per completed referral when accounted for annual inflation rate [13]. These costs are slightly lower than the costs of the m-mama EMTS per referral but four times lower compared to the Tanzania ambulance cost per referral indicating the need for Tanzania to consider scaling up the m-mama EmTS. This difference can be attributed to the difference in health system structure and governance between Tanzania and Burundi and their economic levels

Ambulances are an important part of the emergency transportation systems; however, they have been criticized for being costly and spending on them increases when patients have no other means of transportation in emergency situations [14]. The idea of providing patients with an alternate means of transportation is in line with the findings from the evaluation of the m-mama program presented in this paper demonstrating that an emergency transportation system using community taxis is cost-effective compared to the use of ambulances alone. In the same line, evidence has shown that the use of similar interventions such as *UberX* in emergency services reduced the per capita ambulance volume by at least 6.7% [15].

Cost-effective transportation interventions for obstetric care are an important part of the country’s strategy to reduce maternal mortality because they can address the delay in reaching the health facility which is one of the key drivers of maternal mortality in Low and Middle-Income Countries including Tanzania [16]. Emergency obstetric transportation interventions is more effective when integrated within an enhanced referral system or when additional strategic interventions aimed at improving the quality of care at service delivery points are present [17]. This is an important consideration in scaling up of such interventions they need to be supported by other system improvements such that when a mother goes to the health facility, they get high-quality services they need.

Lack of cost-effectiveness evidence of interventions make their scalability questionable [17]. This study has presented first of a kind cost-effectiveness analysis of an innovative emergency transport system to address maternal mortality in Tanzania. Given the high-cost effectiveness of this intervention compared to the use of ambulances alone, the author recommends this intervention be scaled up in Tanzania in phases with follow-up cost-effectiveness analysis during each phase to generate important evidence for further scale-up and sustainability.

This analysis has several limitations; the data which was complete and eligible to use for the analysis was only for one-year FY2020/2021. The data used in the analysis is about program implementation in only one region and hence regional variability in costs is not considered. Sensitivity analysis of CEA was not done due to lack of enough data and hence the findings limited in generalization.

### Conclusion and Recommendation

The introduction of m-mama emergency transportation system has proven to be cost effective compared to the use of ambulance alone in Shinyanga region. Given the high burden of maternal mortality in Tanzania, this program needs to be scaled up to other regions in Tanzania in a phasic approach and in each phase generating cost-effectiveness analysis upon implementing the intervention in other regions. Moreover, the implementation of the intervention should be accompanied by other health systems strengthening activities like improving the quality of maternal and neonatal care. Importantly, Implementation science methods should be used to document lessons and use the lessons to improve the effectiveness of the EmTS during the scale up phase.

## Data Availability

Since the authors were external evaluators of the implementation of m-Mama program they legally do not own the data, they had limited access during the evaluation period and thus would like to request a waiver on making the data publicly available. Moreover, the data sharing procedure requires signing a Data Transfer Agreement through the National Research and Ethics Committe, a lengthy process that requires amending the consent, which may be difficult since it was not indicated while collecting data, but can always share any analyses required by reviewers or editors.

## Authors’ contribution

Conceptualization: BS, LM, CM, BJN; data cleaning and analysis: GM, LM, BJN, CM; manuscript writing: BS, GM, CM; manuscript revision: BS, LM, JTK, BJN, JEN, NAK, CM, NP, LD, AL

## Acknowledgements

The authors express sincere gratitude to the Directorate of Research and Publications of the Muhimbili University and Allied Sciences (MUHAS) for providing a conducive environment during conceptualization and manuscript writing. Cordial regards to the Touch Foundation, Pathfinder and Vodafone for the innovation and support of the m-mama program. We thank the government officials from the Ministry of Health, President’s Office Regional Administration and Local Government, the Regional and District Health Management Teams for their support in making follows and data validation.

## Funding

This study was funded by Vodafone and Grand Challenge Canada. Both funders had neither role nor influence on the results presented.

## Data Availability Statement

Data will be made available on reasonable request after signing a Data Transfer Agreement (DTA).

## Notes

### Competing Interest Statement

The authors have declared no competing interest.

### Funding Statement

This study was funded by Vodafone and Grand Challenge Canada. LD was the awardee. Both funders had neither role nor influence on the evaluation design, data analysis, decision to publish, preparation of the manuscript and the results presented.

### Author Declarations

The evaluation protocol was submitted to and approved by the Muhimbili University of Health and Allied Sciences (MUHAS) - Institutional Review Board (IRB). Written Informed consent was obtained for participants who participated in the interviews during the evaluation.

